# Barriers to online learning in the time of COVID-19: A national survey of medical students in the Philippines

**DOI:** 10.1101/2020.07.16.20155747

**Authors:** Ronnie E. Baticulon, Nicole Rose I. Alberto, Maria Beatriz C. Baron, Robert Earl C. Mabulay, Lloyd Gabriel T. Rizada, Jinno Jenkin Sy, Christl Jan S. Tiu, Charlie A. Clarion, John Carlo B. Reyes

**Author notes:** **CORRESPONDING AUTHOR:** Ronnie E. Baticulon, MD, Department of Anatomy, University of the Philippines College of Medicine, Manila, Philippines.

## Abstract

**Introduction:** In March 2020, the coronavirus disease 2019 (COVID-19) pandemic forced medical schools in the Philippines to stop face-to-face learning activities and abruptly shift to an online curriculum. This study aimed to identify barriers to online learning from the perspective of medical students in a developing country.

**Methods:** The authors sent out an electronic survey to medical students in the Philippines from 11 to 24 May 2020. Using a combination of multiple choice, Likert scale, and open-ended questions, the following data were obtained: demographics, medical school information, access to technological resources, study habits, living conditions, self-assessment of capacity for and perceived barriers to online learning, and proposed interventions. Descriptive statistics were calculated. Responses were compared between student subgroups using nonparametric tests.

**Results:** Among 3,670 medical students, 3,421 (93%) owned a smartphone and 3,043 (83%) had a laptop or desktop computer. To access online resources, 2,916 (79%) had a postpaid internet subscription while 696 (19%) used prepaid mobile data. Under prevailing conditions, only 1,505 students (41%) considered themselves physically and mentally capable of engaging in online learning. Barriers were classified under five categories: technological, individual, domestic, institutional, and community barriers. Most frequently encountered were difficulty adjusting learning styles, having to perform responsibilities at home, and poor communication between educators and learners.

**Discussion:** Medical students in the Philippines confronted several interrelated barriers as they tried to adapt to online learning. By implementing student-centered interventions, medical schools and educators play a significant role in addressing these challenges during the COVID-19 pandemic and beyond.

## INTRODUCTION

The coronavirus disease 2019 (COVID-19) pandemic has disrupted medical education worldwide [1-6], and the Philippines is no exception. In mid-March, the Philippine government placed its largest island Luzon and other major cities under lockdown, ordering the suspension of classes in all levels [7]. Halfway into the second semester, medical schools had to cease all face-to-face learning activities and abruptly transition to various forms of remote or online learning. Whether the original learning outcomes could be attained, or if it is reasonable to expect such in a pandemic situation, has remained unclear. There are no studies that describe the utilization of and current capacity for online learning in Philippine medical schools. This is complicated further by marked variability in medical curricula throughout the country [8].

The Doctor of Medicine program in the Philippines takes four years. The first two years consist mainly of didactic teaching, and the final year is clerkship, when medical students must complete clinical rotations in internal medicine, pediatrics, surgery, and obstetrics-gynecology [9]. Afterward, to obtain a medical license, students must undergo one year of internship and then pass a written examination that covers 12 subjects. The pandemic has derailed all of these, with medical students pulled out of their clinical rotations, the national internship program suspended, and the licensure exam postponed indefinitely.

At the time of this writing (July 10), the Philippines has documented 51,754 confirmed cases of COVID-19 [10]. As infections and deaths continue to rise, medical schools only have several weeks to restructure their curriculum, train faculty, and prepare students for the next academic year. Face-to-face classes and hospital rotations are not expected to resume until September, at the earliest.

Previously published studies have focused on COVID-19’s impact on medical education in high-income countries [2,11-13]. Data in developing countries are sparse and urgently needed [14-17]. In this paper, we aimed to identify and describe the challenges to online learning—also called e-learning, web-based learning, or internet-based learning [18,19]—among medical students in the Philippines during the COVID-19 pandemic. We hypothesized that students have faced socioeconomic and cultural barriers, in addition to limited access to technological resources. It is crucial to address these barriers in low- and middle-income countries, where the need to continuously train skilled health workers is also greatest [20].

## METHODS

### Setting and participants

We conducted a nationwide cross-sectional study among medical students in the Philippines from May 11 to 24, 2020 through an electronic survey in Google Forms (Google LLC, Mountain View, California). We distributed the survey link through social media (Twitter, Facebook, Instagram) and the Association of Philippine Medical Colleges Student Network. We contacted student organizations to share the link among students in their respective schools. The survey was open to students from first to fourth year level. Participation was voluntary, anonymity was guaranteed, and consent was obtained at the start of the survey. The University of the Philippines Manila Research Ethics Board reviewed the study protocol and issued a certificate of exemption (UPMREB 2020-281-EX).

### Survey instrument

We initially conducted a focus group discussion with medical students from the University of the Philippines, reviewed relevant literature, and searched official school websites for announcements on changes being implemented in Philippine medical schools due to the COVID-19 crisis [21-24]. We looked for social media posts of medical students and their organizations in reaction to these curricular adjustments [25]. Using these background data, we developed a 23-item questionnaire that collected demographics, medical school information, access to technological resources, study habits, current living conditions, and views on online learning. We used Howlett’s definition of online learning, which is ‘the use of electronic technology and media to deliver, support, and enhance both learning and teaching and involves communication between learners and teachers utilizing online content [26].’

Using a 4-point Likert scale (strongly disagree, disagree somewhat, agree somewhat, strongly agree), we asked the participants: (1) whether they considered themselves physically and mentally capable of studying online for the rest of the semester; (2) whether medical schools should give a passing grade to all their students (i.e., mass promotion); (3) whether they had enough time and resources for online learning; and (4, 5) whether the resources of their schools and the skills of their educators were adequate. We listed ten barriers to online learning and asked respondents to select how frequently they have encountered each barrier (never, sometimes, often, always). In open-ended questions, we probed for any additional barriers that the students may have faced, and asked for their proposed interventions.

### Data analysis

De-identified data were exported to Stata/IC 16.1 for Mac (Stata Corp, College Station, Texas). We stratified the data based on demographics and medical school information, calculating the frequencies and percentages of categorical variables. In the self-assessment of capacity for online learning, the responses were converted to their numeric equivalents from 1 to 4, and these were used to get the mean and median responses for each subgroup. We analyzed differences between subgroups using Kruskal-Wallis test, with post hoc analysis using Dunn’s multiple nonparametric pairwise tests. A *P* value < 0.05 was considered statistically significant. For the rest of the Likert scale questions and the list of barriers to online learning, we computed the frequencies and percentages of responses. Finally, we performed thematic analysis on the answers to open-ended questions.

## RESULTS

We received a total of 3,813 responses. We removed 75 (2%) that were duplicate entries based on email addresses or student numbers, and 68 (2%) that were deemed invalid because of missing information or because respondents were not medical students. Hence, we included 3,670 responses (96%) in the data analysis. This represented 15% of the estimated 25,000 medical students in the Philippines.

The exact number and distribution of medical students enrolled for academic year 2019–2020 were not available at the time of study completion. In lieu of this, we compared our nonrandom study sample with the total population of first-time examinees of the physician licensure exam in the last five years [27] (See Table 1). Our sample had a higher percentage of students enrolled in public medical schools (16% vs. 14%, p = 0.001). The representation of the different regions also differed (p < 0.001). In particular, the proportion of medical students from the National Capital Region was lower (47% vs. 55%) and that from Mindanao was higher (13% vs 7%) in our study. Of the 55 medical schools recognized by the Commission on Higher Education, 54 (98%) were represented in this survey. The median number of respondents per institution was 42 (range: 1–293), with 23 schools having 50 respondents or more.

**Table 1:** Comparison of study population with first-time examinees of the national physician licensure examination from August 2014 to March 2019

| Medical School Category |  | Current Study |  | Board Examinees, 2014 to 2019* |  | P value |
| --- | --- | --- | --- | --- | --- | --- |
|  |  | Number of Students (N = 3,670) | Percentage | Number of Students (N = 19,124) | Percentage |  |
| **Classification** | Public | 592 | 16% | 2,682 | 14% | 0.001 |
|  | Private | 3,078 | 84% | 16,442 | 86% |  |
| **Location** | National Capital Region | 1,726 | 47% | 10,492 | 55% | < 0.001 |
|  | Luzon | 728 | 20% | 3,276 | 17% |  |
|  | Visayas | 721 | 20% | 3,928 | 21% |  |
|  | Mindanao | 495 | 13% | 1,428 | 7% |  |

\*Source: Professional Regulation Commission

### Demographics and access to technological resources

More students from lower year levels answered the survey: 1,153 (31%), first year; 1,015 (28%), second year; 863 (24%), third year; and 639 (17%), fourth year. Mean age was 23.8 ± 2.4 years. At a ratio of 2.2:1, females (n=2,468, 67%) outnumbered males (n=1,109, 30%). There were 39 (1%) who identified as non-binary. Among the respondents, 169 (5%) were married or partnered in a long-term relationship while 82 (2%) had children. The majority of medical students belonged to the low-income and lower-middle-income brackets (n=1,029, 28% and n=1,665, 45%, respectively). One out of six (n=651, 18%) received financial aid.

On device ownership, 3,421 (93%) had a smartphone, 2,311 (63%) had a tablet, and 3,043 (83%) had a laptop or desktop computer. There were 216 students (6%) who owned a smartphone only. Most students (n=2,916, 79%) subscribed to a postpaid internet service, but three out of five subscribers described their connection as slow and/or unreliable. Accessing the internet primarily through prepaid mobile data was still common at 19% (n=696). While most students lived at home with their family during the pandemic (n=2,856, 78%), there were 610 (17%) who remained in their temporary residence near their school.

### Capacity for online learning

Under prevailing circumstances, only 41% of medical students (n=1,505) considered themselves capable of adapting to online learning. The responses of the students were similar, regardless of medical school classification (p = 0.79) or location (p = 0.96) (See Table 2). Factors that significantly affected the self-assessment of students were year level (p < 0.001), age (p = 0.002), gender (p < 0.001), annual family income (p < 0.001), academic standing (p < 0.001), internet access (p = 0.03) and number of hours previously spent on online learning (p < 0.001). Students less likely to consider themselves capable of online learning were in their first or second year of medical school, 29 years old or younger, female or non-binary, or from a family in a low- or middle-income bracket. Negative responses were also more common among those who reported a lower academic standing or previously spent fewer hours on online learning every week.

**Table 2:** Comparison of self-assessment of capacity for online learning among student subgroups*

| Category | Subgroup | Number of Students and Percentage (N = 3670) |  | Capacity for Online Learning* |  | P value |
| --- | --- | --- | --- | --- | --- | --- |
|  |  |  |  | Mean (SD) | Median |  |
| **Medical school location** | National Capital Region | 1,726 | 47% | 2.28 (0.83) | 2 | 0.792 |
|  | Luzon, Visayas, and Mindanao | 1,944 | 53% | 2.29 (0.86) | 2 |  |
| **Medical school classification** | Public | 592 | 16% | 2.29 (0.84) | 2 | 0.958 |
|  | Private | 3,078 | 84% | 2.29 (0.85) | 2 |  |
| **Year level** | 1st and 2nd year | 2,168 | 59% | 2.21 (0.84) | 2 | < 0.001 |
|  | 3rd and 4th year | 1,502 | 41% | 2.41 (0.85) | 2 |  |
| **Age** | < 30 years old | 3,570 | 97% | 2.28 (0.85) | 2 | 0.002 |
|  | ≥ 30 years old | 100 | 3% | 2.58 (0.87) | 3 |  |
| **Gender†** | Male | 1,109 | 30% | 2.45 (0.86) | 2 | < 0.001 |
|  | Female | 2,468 | 67% | 2.22 (0.84) | 2 |  |
|  | Non-binary/Prefer not to say | 93 | 3% | 2.25 (0.79) | 2 |  |
| **Family relationships** | Married/partnered or with children | 201 | 5% | 2.39 (0.84) | 2 | 0.086 |
|  | Rest | 3,467 | 94% | 2.28 (0.85) | 2 |  |
| **Family income status‡** | Low income | 1,029 | 28% | 2.23 (0.86) | 2 | < 0.001 |
|  | Lower- and upper-middle income | 2,306 | 63% | 2.29 (0.83) | 2 |  |
|  | High income | 302 | 8% | 2.47 (0.95) | 2 |  |
| **Self-reported academic standing‡** | Highest 25% in year level | 568 | 15% | 2.55 (0.90) | 3 | < 0.001 |
|  | Middle 50% in year level | 2,581 | 70% | 2.27 (0.83) | 2 |  |
|  | Lowest 25% in year level | 521 | 14% | 2.09 (0.82) | 2 |  |
| **Device ownership\*\*** | One device or none | 375 | 10% | 2.25 (0.93) | 2 | 0.167 |
|  | Two devices or more | 3,295 | 90% | 2.29 (0.84) | 2 |  |
| **Internet access** | No access | 58 | 2% | 2.07 (0.95) | 2 | 0.030 |
|  | Prepaid mobile data | 696 | 19% | 2.25 (0.87) | 2 |  |
|  | Postpaid subscription | 2,916 | 79% | 2.30 (0.84) | 2 |  |
| **Hours spent on online learning, prior to pandemic‡** | 4 hours/week or less | 1,552 | 42% | 2.21 (0.85) | 2 | < 0.001 |
|  | 5 to 24 hours/week | 1,848 | 50% | 2.33 (0.83) | 2 |  |
|  | 25 to 40 hours/week | 270 | 7% | 2.47 (0.94) | 3 |  |
| **Hours allocated in school schedule for independent study, prior to pandemic** | 4 hours/week or less | 1,620 | 44% | 2.28 (0.85) | 2 | 0.526 |
|  | 5 to 24 hours/week | 1,689 | 46% | 2.30 (0.83) | 2 |  |
|  | 25 to 40 hours/week | 361 | 10% | 2.28 (0.93) | 2 |  |

\* The respondents were asked to indicate whether they perceived themselves to be “physically and mentally capable of studying all remaining subjects for the semester online.” Possible responses were strongly disagree (1), disagree somewhat (2), agree somewhat (3), and strongly agree (4). Numeric equivalents were used to compute for mean and median responses for each subgroup. The means and standard deviation are indicated for ease of comparison, but *P* values were obtained using nonparametric tests.
† On post hoc analysis, males were significantly different from the rest.
‡ On post hoc analysis, all subgroups were significantly different from each other.
§ Annual income brackets: low income, Php 250,000 or less; lower-middle income, between Php 250,000 and Php 1 million; upper-middle income, between Php 1 and 2 million; high income, Php 2 million or more. Php 50 : US\$1.
\*\* Device may be a smartphone, tablet, laptop, or desktop computer.

Figure 1 summarizes the responses to the Likert scale questions. The majority of the respondents (n=2,651, 72%) believed that medical schools affected by the COVID-19 pandemic should promote all students, with 77% (n=2,813) saying that they had enough time and resources to prepare for the next year level. Almost half (n=1,720, 47%) agreed that their teachers had the requisite skills and resources, while 44% (n=1,604) said that their schools were equipped to support online teaching.

**Figure 1:**
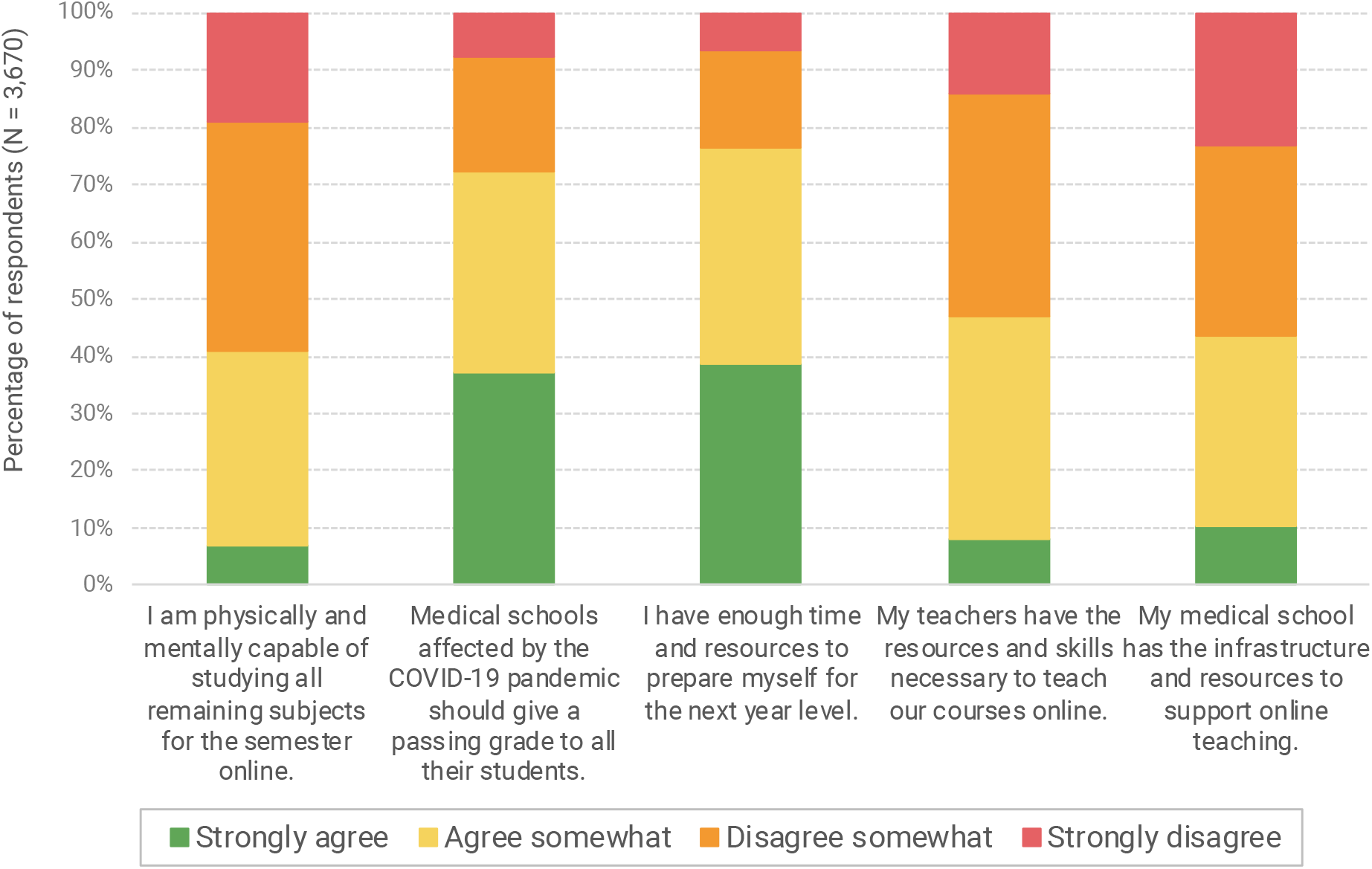
Summary of responses to Likert scale questions.

### Barriers to online learning

Among barriers to online learning, the most frequently encountered were difficulty adjusting learning styles, having to perform responsibilities at home, and poor communication or lack of clear directions from educators (See Figure 2). Approximately two-thirds of the respondents always or often confronted these barriers. Lack of physical space conducive for studying and mental health difficulties were also common. The data showed that the availability of fast and reliable internet connection was a bigger concern than either device ownership or technical aptitude. One out of ten students always or often lacked basic needs such as food, water, medicine, and security. Internal consistency for these ten barriers showed acceptable reliability with Cronbach’s α = 0.78.

**Figure 2:**
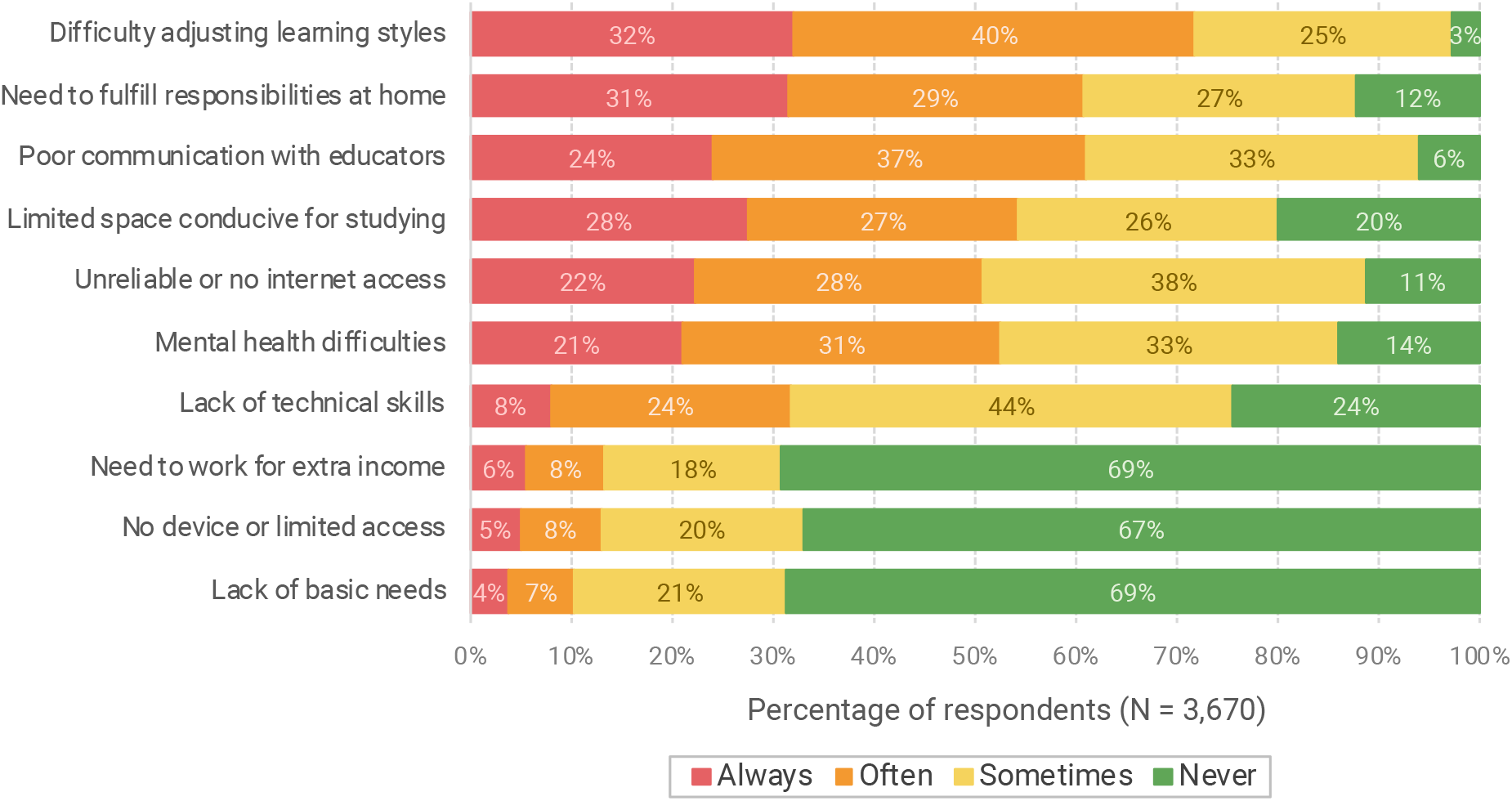
Frequency of occurrence of selected barriers to online learning among medical students in the Philippines during the COVID-19 pandemic.

We elicited additional challenges to online learning in the free-text responses. These are summarized in Table 3. Together with our initial list, the barriers were grouped under five categories: (1) technological, which pertain to hardware, software, and internet connectivity; (2) individual, which involve students’ learning styles, physical, and mental health; (3) domestic, which are concerns at home or within the family, including financial distress; (4) institutional, which revolve around administration, medical curriculum, school resources, and educator skills; and (5) community barriers, which include lockdown restrictions, infrastructure challenges, and sociopolitical issues.

**Table 3:** Summary of student barriers to online learning during the COVID-19 pandemic

| CATEGORY | BARRIERS | SAMPLE RESPONSES FROM STUDENTS |
| --- | --- | --- |
| Technological barriers | • Lack of devices or limited access due to gadget sharing<br>• Unreliable, slow, or no internet access<br>• Lack of technical skills<br>• Issues with the online learning platform | • “We only have one desktop computer, which is also used by my brother, a student himself.”<br>• “I live outside the city where my school is located. The internet connection is poor and unreliable.”<br>• “I only use mobile data and internet credit is expensive.”<br>• “Our school uses [REDACTED] as the main platform for online learning. It crashes often.” |
| Individual barriers | • Difficulty adjusting learning styles<br>• Mental health difficulties<br>• Physical health issues<br>• Practical concerns | • “Having a hard time understanding materials on my own.”<br>• “Lack of drive to study since it’s different from the school setup.”<br>• “Procrastination, distractions like unlimited internet access.”<br>• “This pandemic gives nothing but uncertainty, stress, and anxiety.”<br>• “Eye strain and headache from prolonged use of gadgets.”<br>• “My books, reference materials, and printer were all left in my boarding house.” |
| Domestic barriers | • Limited space conducive for studying<br>• Need to fulfill responsibilities at home<br>• Conflicts within the family<br>• Financial distress within the household<br>• Need to work for extra income<br>• Lack of basic needs | • “[Home] is not conducive [for studying] because of small space and noisy background.”<br>• “I need to allocate a whole day just to buy groceries, medicine, and other supplies because of the exhausting lines in each.”<br>• “Relationship with family members is strained, so being in the house for so long is emotionally and mentally tiring.”<br>• “I have to work for extra income since the main source of income of our family is affected by the pandemic.”<br>• “We had no choice but to subscribe to an internet service provider, despite my family being on a tight budget.” |
| Institutional barriers | • Administrative issues and lack of organization<br>• Poor communication between learners and educators<br>• Inadequate skills of educators<br>• Poor quality of learning materials<br>• Gaps in knowledge and skills from current teaching methods<br>• Excessive cognitive load<br>• Limited opportunities to interact with peers<br>• Policies and practices that neglect student welfare | • “Our school has not officially provided us with plans should the current situation persist.”<br>• “We were promised leniency but our voices aren’t heard.”<br>• “[Teachers] keep saying that they want to assess us but then they never give feedback to the students.”<br>• “Lack of preparedness of educators to shift to online learning.”<br>• “Some professors just gave their files [presentation slides], with no audio or presenter notes.”<br>• “Insufficient patient exposure for practical learning. In my case, skills in missed clinical rotations such as orthopedics and radiology.”<br>• “Workload is far too much compared to when there were face-to-face classes.”<br>• “[There is] the need for peers, for social connection and motivation.”<br>• “My school requires two gadgets for them to watch on the other gadget while I’m taking the exam on another device.” |
| Community barriers | • Mobility restrictions due to community lockdown<br>• Power interruptions<br>• Sociopolitical concerns | • “Curfew hours [affect me because] I take my exams at my friend’s house for better internet connection.”<br>• “Rotational brownouts in provinces lasting for about 2 to 6 hours.”<br>• “As a concerned Filipino, with all that’s happening in the government, it is so hard to just hunch over these books without being distracted.” |

## DISCUSSION

This national survey of 3,670 medical students from 54 schools in the Philippines revealed that students, regardless of geographic location or demographic subgroup, have encountered several barriers as they tried to adapt to online learning during the COVID-19 pandemic. Gender, age, year level, annual income, academic standing, internet access, and number of hours previously spent on online learning affected the medical students’ perception of their capacity to engage in online learning. We classified the barriers under five categories: technological, individual, domestic, institutional, and community barriers.

The advantages of using online learning in medical education include improved accessibility of information, ease of standardizing and updating content, cost-effectiveness, accountability, and enhancement of the learning process, wherein students are motivated to be active learners [19,28]. It has been shown to be equivalent, possibly even superior to traditional methods of curriculum delivery [29]. In low- and middle-income countries, it has the potential to address faculty shortage, expanding the reach of medical educators and improving their efficiency [23]. Despite these, in recent single-center studies in India and Pakistan, the majority of medical students had a negative perception towards online learning [14,30]. A similar survey in Indonesia showed that many were concerned about lack of interaction, and difficulty concentrating and understanding concepts online [16].

A significant limitation common to these studies was that the participants belonged to a single medical school. This mirrored the findings of Barteit et al. in their review of studies that evaluated e-learning interventions for medical education in developing countries [22]. They found that most studies were small-scale and had examined projects in their pilot stages; this phenomenon coined ‘pilotitis’ has hindered the development of standards for e-learning in low-resource settings. Further, research in developing countries has focused mostly on technological or contextual challenges, often failing to provide a comprehensive view or whole-system perspective [21]. It is important to identify any additional enablers and barriers, which may not have been present in the high-income countries where these teaching strategies were often developed and first evaluated.

Before the COVID-19 pandemic, medical schools in the Philippines have never had to implement online learning on this massive scale. The Commission on Higher Education’s guidelines on the Doctor of Medicine program have not set standards and minimum resource requirements for remote or online learning [9,31]. This unprecedented situation presents an opportunity to critically examine the state of medical education nationwide, systematically evaluate the effectiveness of online curricula in a developing country, and formulate contingency plans for similar circumstances in the future. Our paper provides important baseline data for these efforts.

We have shown that the number of medical students with limited access to technological resources is not negligible. One out of five students did not have a computer, and an identical proportion had to rely on prepaid mobile data for connectivity. Roughly one out of twenty used only a smartphone. Power interruptions, weak infrastructure, and internet costs restricted the students’ access to online content, similar to other developing countries [16,32-34]. The striking finding is that students did not perceive these technological limitations to be the most important barriers, as can be seen in Figure 2. This suggests that students have somehow managed to cope with these challenges during the acute phase of the pandemic. It also implies that providing gadgets to the students, as some medical schools had already done, may not be enough to ensure successful learning outcomes.

Our data confirmed that traditional teaching methods (i.e., teacher-led, classroom-based learning activities) continue to be the norm in Philippine medical education. Almost three-fourths of the respondents (74%) indicated that prior to the pandemic, no more than eight hours were allocated to self-directed learning in their weekly schedules. Consequently, the abrupt shift in curriculum delivery, which required a simultaneous adjustment in learning styles, had been difficult for the students. Those who previously spent fewer hours studying online were less likely to agree that they could cope. Respondents said that they needed more time to comprehend learning materials. Many admitted that they lacked self-discipline and drive to study. Educators must understand learners’ needs, motivations, and past experiences to maintain engagement in an online curriculum [28]. To achieve academic success, students need to be guided in developing self-regulated learning strategies, which include time management, metacognition, critical thinking, and effort regulation [35].

The pandemic had also caused psychological stress among the students, making it difficult for them to focus on studying. They expressed feelings of anxiety, burnout, loneliness, homesickness, grief, and hopelessness. The students worried about online assessments, future plans in medical school, possible delays in training, and safety of their families from COVID-19. Overall, 86% of the students reported experiencing some degree of mental health difficulty. We noted that difficulty adjusting learning styles and mental health concerns were more common among female and non-binary respondents, those in the first two years of medical school, and those with a lower academic standing. This may partly account for observed differences in self-assessment of capacity for online learning. Further studies to elaborate on these factors are warranted. Living in urban areas, economic stability, and living with parents have been shown to be protective factors against anxiety among students in China [36].

It was evident that more time spent at home did not necessarily equate to more time for academic work. There were students who could not concentrate because they were constantly exposed to conflict among family members. Even in the absence of domestic dispute, some found it hard to turn down conversations with parents or siblings. Filipino families are characterized by cohesiveness and reciprocity, and the most educated members are often expected to act as caregivers or household heads [37-39]. In the current health crisis, many medical students had been relegated to this role. They took care of sick relatives, were in charge of buying food and supplies, or had to work for extra income. Moreover, although the learning environment may be virtual, physical space remained vital. Having a quiet study area, with the same comfort provided by a classroom or library, was a privilege not available to all.

Medical students doubted the readiness of their schools to transition to online learning. They cited lack of guidelines, unfair policies, haphazard class schedules, low quality of teaching materials, ineffective teaching strategies, and excessive class requirements. For comparison, academic medical centers in Singapore have clearly laid out allowable undergraduate education activities and assessments depending on their pandemic alert level [11]. Communication channels needed improvement. Students said that their views were not being heard, and they lamented the lack of appropriate action on their feedback. Students were concerned that they were not learning essential skills or getting ample patient exposure, a sentiment that is echoed around the globe [40,41]. They also voiced out the need to interact with peers, with whom they could exchange insights, resources, and opinions.

The current shifts in Philippine medical education have been happening with the economic consequences of COVID-19 in the background. Unemployment rate in the country is at record high [42]. Analysis of the students’ responses had revealed the pervasive nature of this problem: working students lost jobs that supported their education; household budgets had to be split between essential needs and internet subscription; family-owned businesses closed; and scholarships were put in limbo. Consistent with these responses, students from lower income brackets felt less capable of engaging in online learning. Some expressed that they might not be able to enroll next school year because they would not be able to afford the cost of medical education.

Almost three-fourths of the respondents belonged to families with an annual income of less than one million pesos, yet only one out of six declared being the beneficiary of a financial scholarship. Tuition alone ranges from 24,000 to 150,000 pesos per semester [43]. The added cost of online learning should not be underestimated. An hour of video lectures will consume about 480 MB of mobile data [44]. At the prevailing rate of approximately 23 pesos per GB, a student who watches four hours of videos will need to spend 45 pesos daily. To put the figures in context, minimum daily wage in the Philippines in May 2020 ranged from 230 to 450 pesos [45].

Our framework emphasizes that the challenges to online learning in developing countries are multifactorial and interrelated, especially during a global health crisis. With these study findings, after review of proposed interventions from the respondents, we put forward the following recommendations for medical schools:

1. Conduct a needs assessment survey among medical students to identify those with limited access to technological resources and basic needs.
2. Ensure open communication channels among administrators, educators, and students (e.g., through online town hall meetings). Guidelines and expectations must be clear, with provisions for improvement or worsening of the pandemic situation.
3. Implement a primarily asynchronous mode of content delivery with minimal technical and data requirements. Smartphone compatibility remains essential.
4. During the transition phase, maximize use of curated online resources that are available for free or with an institutional subscription. To sustain the online curriculum, support and train faculty on content creation, management, and delivery. Invest in technical support.
5. Avoid cognitive overload. As with classroom teaching, ensure that assessment measures are aligned with desired learning outcomes.
6. Extend leniency to students who bear additional responsibilities at home.
7. Develop mental wellness programs and provide proactive psychosocial support for the students.
8. Create opportunities for meaningful interaction with peers and mentors.
9. Give discounts on tuition and offer scholarships to cushion the pandemic’s economic impact. Advocate for greater subsidies from the government.
10. Develop bridging programs and prepare for gradual return to clinical activities. Consider putting up simulation laboratories and other infrastructure that will allow face-to-face learning with social distancing. Look for sustainable means to provide personal protective equipment for students.

Our study was subject to selection bias, wherein students with no internet access and those who had been severely affected by the pandemic may not have received our survey. Social distancing measures had already been in place during the study period, preventing the distribution of questionnaires in person. Thus, the reported deficit in technological resources is likely an underestimate. Self-reporting bias may have also affected the students’ responses.

Some of the barriers we have enumerated are transient and expected to resolve with the global health crisis. Others may persist, or have long-term consequences. Without appropriate intervention, these barriers would not just affect the education and training of future physicians. On a wider scale, even the nation’s delivery of health care services may be disrupted. It has become apparent that many of these barriers were pre-existing, with disparities between subgroups being heightened by the pandemic, often in favor of those with greater access to resources. This leads to an unequal learning environment, albeit unintended, and the greatest challenge for educators is to ensure that this inequity is not perpetuated.

## Data Availability

All data are secured by the primary investigator. Requests for data should be directed to the primary investigator, subject to the guidelines of the University of the Philippines Manila Research Ethics Board.

## ACKNOWLEDGEMENTS

The authors would like to acknowledge the assistance of the Commission on Higher Education’s Office of Programs and Standards Development, and the support of the Association of Philippine Medical Colleges–Student Network, the Asian Medical Students’ Association–Philippines, and the student councils of the following medical schools: Ago Medical and Educational Center–Bicol Christian College of Medicine, Ateneo De Zamboanga University, Bicol University, Cagayan State University Carig, Cebu Doctors’ University, Cebu Institute of Medicine, Cebu Normal University, De La Salle Medical and Health Sciences Institute, Remedios T. Romualdez Medical Foundation, San Beda University, Silliman University, University of the Philippines Manila, West Visayas State University, and Xavier University.

